# Assessment of Individual- and Community-level Risks for COVID-19 Mortality in the US and Implications for Vaccine Distribution

**DOI:** 10.1101/2020.05.27.20115170

**Authors:** Jin Jin, Neha Agarwala, Prosenjit Kundu, Benjamin Harvey, Yuqi Zhang, Eliza Wallace, Nilanjan Chatterjee

**Affiliations:** Department of Biostatistics, Bloomberg School of Public Health, Johns Hopkins University; Department of Mathematics and Statistics, University of Maryland, Baltimore County; Department of Biomedical Engineering, Johns Hopkins University; PolicyMap Corp, Philadelphia, PA; Department of Oncology, School of Medicine, Johns Hopkins University

## Abstract

Reducing COVID-19 illness and mortality for populations in the future will require equitable and effective risk-based allocations of scarce preventive resources, including early available vaccines. To aid in this effort, we develop a risk calculator for COVID-19 mortality based on various socio-demographic factors and pre-existing conditions for the US adult population by combining information from the UK-based OpenSAFELY study, with mortality rates by age and ethnicity available across US states. We tailor the tool to produce absolute risks for individuals in future time frames by incorporating information on pandemic dynamics at the community level as available from forecasting models. We apply this risk calculation model to available data on prevalence and co-occurrences of the risk-factors from a variety of data sources to project risk for the general adult population across 477 US cities (defined as Census Places) and for the 65 years and older Medicare population across 3,113 US counties, respectively. Validation analyses based on these projected risks and data on tens of thousands of recent deaths show that the model is well calibrated for the US population. Projections show that the model can identify relatively small fractions of the population (e.g. 4.3%) which will lead to a disproportionately large number of deaths (e.g. 49.8%), and thus will be useful for effectively targeting individuals for early vaccinations, but there will be wide variation in risk distribution across US communities. We provide a web-based tool for individualized risk calculations and interactive maps for viewing the city-, county- and state-level risk projections.

## Introduction

The first case of SARS-CoV-2 infection in the US was reported on January 20^th^, 2020, in the state of Washington,^1^ and to date the pandemic has led to more than 168,000 COVID-19 deaths - making the US by far the most affected country globally. However, there is a major variation in rates of infections and underlying deaths across US states, counties and cities. Various local population characteristics, such as mitigation measures,^2,3^ population density and mobility patterns^4,5^ define background risks of illness and death across the regions. Further, epidemiologic studies have provided evidence that a variety of pre-disposing factors, including age, gender, ethnic and racial background, social conditions and pre-existing conditions, put individuals within the same community at differential risks of serious illness and mortality.^6-14^

To date, the US and other countries have mostly relied on community-based intervention measures, such as lockdowns, social distancing, and guidance on mask wearing, for mitigating the worst effects of the pandemic. A variety of pandemic scenario models are available for forecasting future trends in infection, hospitalizations, and deaths at the population level. Although the existence of a variety of predisposing factors has been known, there has been limited effort to incorporate these factors into prevention strategies or/and forecasting models. In the future, however, as the US and other countries continue to face increasing societal and economic pressure for relaxing some of the broad intervention measures, consideration of risk associated with predisposing factors at the individual- and population-level will be important in developing more equitable strategies for prevention.^15-17^ Specifically, as promising results from early phases of a number of vaccines trials^18,19^ have raised the likelihood of available vaccines within a few months, there has been a recognition that a framework is urgently needed for equitable distribution of the limited supply of early vaccines. Recently, the US National Academies of Science and Engineering announced the launch of a study to develop criteria for early vaccination based on various individual- and community-level risk information.^20^ Further, until vaccination is possible, risks calculations can also be critical for identifying high-risk groups who should be prioritized for “shielding”^16,17^ as the pandemic continues and yet various socioeconomic activities resume.

In this article, we describe the development and validation of a COVID-19 mortality risk calculator for the US adult (18+ year old) population integrating information from a variety of datasets for estimation of risk associated with predisposing factors. Using a first of its kind method, we further extend the calculator to integrate information from pandemic forecasting models so that an individual’s absolute risk can be informed based not only on their underlying risk-factors, but also on community-level risk due to the underlying pandemic dynamics. We use the information on the prevalence and co-occurrence of risk-factors from various national databases to make population-level projections of risks associated with these predisposing factors for the general adult population across 477 US cities, and for the 65+ year old population enrolled in Medicare across 3,113 US counties. We provide country-, state- and city/county- level estimates for the size of populations who are at or above different risk-thresholds and thus can be gradually prioritized for vaccination and other preventive efforts. Finally, we provide a web-based individual-level risk calculator and interactive maps for viewing the population-level risk projections to facilitate future policy decisions.

## Methods

### Definition of COVID-19 mortality risk-score

The risk-score for an individual is defined as a weighted combination of various socio-demographic characteristics and predisposing health conditions, with weights defined by the relative magnitude of the contribution of these factors to the risk of death due to COVID-19 in the adult population. We use two sources of information to build the risk-score: (1) multivariate-adjusted estimate of risk associated with gender, social deprivation index and 12 pre-existing conditions from the recently published UK-based OpenSAFELY study,^13^ and (2) death rates associated with different age and racial/ethnic groups in the US published by the Center of Disease Control,^21^ after performing external covariate adjustment accounting for the correlation of these factors with other risk factors in the model (See Below).

### Estimation of US-specific risk associated with age and racial/ethnic groups and external covariate adjustments

We used data made available by the CDC^21^ on reported COVID-19 deaths as of June 6, 2020, by race, age, and state. We fitted a Poisson regression to the death counts available by age, race, and state with underlying population sizes as offset terms for modeling rates. We modelled the lograte in terms of additive effects of age and race/ethnicity categories and further adjusted for states as fixed effects in the model. We then used data available on age, race/ethnicity and all other risk factors of the model from a combination of health survey data (see below) to estimate joint distribution of all of these factors, and use Generalized Method of Moment (GMM) techniques we have developed earlier^22^ to obtain estimates for the effects associated with age and race adjusted for the other risk-factors in the model with their effects being fixed at those available from an underlying fully adjusted model from the UK OpenSAFELY study (Supplementary Methods Section 1.2, Extended Data Table 1).

### Data sources for obtaining prevalence and joint distribution of the risk factors in the general adult population

We utilized a variety of data sources to obtain the latest information on the prevalence of demographic variables and health conditions across the US cities. The resources include American Community Survey (ACS) of the US Census Bureau for age, gender and race (2017/2018 table, one-year estimates),^23^ Behavioral Risk Factor Surveillance System (BRFSS, 2017 survey) of the Centers for Disease Control and Prevention (CDC) for various health conditions and smoking,^24^ National Health and Nutrition Examination Survey (NHANES) for estimating relative proportions of certain sub-categories of conditions that were not available in BRFSS,^25^ United States Cancer Statistics (2012-2016 incidence rates) maintained by the National Cancer Institute and the CDC to derive prevalence of hematological and non-hematological malignancies,^26^ and a database available from the Robert Graham Center on social deprivation index (SDI) derived from data available from the ACS (2011-2015 five-year estimates).^27^ Detailed derivations of each variable are described in the Supplementary Methods, Section 1.1. In addition, we accessed individual-level data from the National Health Interview Survey (NHIS) of CDC.^28^ We extracted individual-level data on the risk factors on 22,109 adults from the 2017 NHIS study. All of the required variables, except SDI, were available for individuals in the NHIS. For projections of risk for the US overall, we applied the most recent age distribution from the US Census Bureau 2019 data,^29^ and information of the other risk-factors within age groups from the NHIS.

### US Medicare population

We used 2018 data from the Centers for Medicare and Medicaid Services (CMS) to obtain the latest information on prevalence of chronic health conditions across US counties for the 65+ year old Medicare population (the <65 year old Medicare population have missing information on age and thus were not included in the analyses).^30^ We utilized the data for 65+ year old individuals from 2017-18 NHANES^25^ and 2017 NHIS^28^ to estimate relative proportions of certain subcategories of the health conditions that were not available from the CMS. Prevalence of race and gender variables for the Medicare population were obtained from the ACS.^31^

### Statistical models and methods

Similar to the OpenSAFELY study, we assume that the risk of COVID-19 death at time *t* for an individual *i* residing in location *l*, e.g. a city or a county, can be described by the proportional risk model

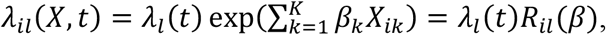

where *λ_l_*(*t*) denotes the baseline risk for location *l* due to the underlying pandemic characteristics, and 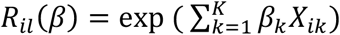 denotes a multiplicative factor associated with risks due to various predisposing factors. Here *t* refers to calendar time since some landmark, such as the day when the cumulative death reaches some minimum threshold. The average risk of the population at location *l* can be defined as

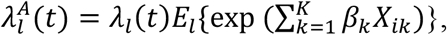

where *E_l_* denotes the expectation (average) with respect to distribution of the risk factors in location *l*. The above formula allows linking individual-level relative risk models to pandemic scenario models and hence can produce estimates of absolute risk of individuals taking into account both individual-level risk-factors and community-level risk due to pandemic dynamics. In particular, a variety of pandemic models are available to produce estimates of population level risk 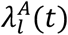, e.g in the state of residence of an individual, over the course of a period of time in the future, and such information can be used to calculate the baseline risk *λ_l_*(*t*) and hence the absolute risk denoted as *λ_il_*(*X, t*) (Supplementary Methods, Section 2.1).

We define the quantity 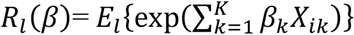 as an Index of Excess Risk (IER) for the population associated with the underlying risk-factor distribution in location *l*, and present the scaled version of IER as 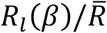, where 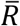 denotes the weighted average of *R_l_*(*β*) across cities/counties with population sizes as the weights. Further, we examine the distribution of *R_il_*(*β*) across individuals within a location to identify the size of the underlying most “vulnerable” populations. For these evaluations, ideally one would require individual-level data for a representative sample of individuals from each location. However, in the absence of such data, we develop a framework to approximate the distributions using city/county-specific information on prevalence, and information on the co-occurrence of these factors, captured through the underlying odds-ratio parameters, from NHIS. Further, we assume a normal or mixture normal distribution of the underlying risk-scores within each location and use the individual-level data available from NHIS to evaluate the accuracy of the approximations for tail probability calculations (Extended Data Figure 1, Supplementary Methods Section 2).

### Model validation

We conduct an independent validation of the risk projections using county-level mortality information from the CDC between June 7 and August 1, which did not contribute to the model development. Specifically, for each of the 256 counties that contain the 477 studied cities, we calculate a weighted IER with each city weighted by its population size. We then examine how strongly the county-level IER predicts the underlying death rates using two approaches. First, we fit a negative binomial model where the log of death rates across the counties are modelled as a linear function of log(IER), and residual heterogeneity in the model is accounted for using a Poisson-Gamma random effects model. If the underlying individual-level risk model is correctly specified, then in this group-level model, one would expect the slope of log(IER) to be close to 1.0. We further use weighted least squares to estimate a measure of explained variance (*R*^2^) of log(death rate) associated with log(IER). As a benchmark, we also estimate similar measures for two other likely predictors, log of population density^32^ and log of three-week prior infection rate.^33^ We also conduct a conditional analysis to account for major regional differences in pandemic dynamics between the Northeast, Midwest, South, and West.^34^ All analyses are done using information on deaths over a moving window of two-week periods for the detection of potential temporal effects. Details can be found in Supplementary Methods, Section 2.5.

## Results

Risks of mortality associated with various age groups in the US follow a very comparable pattern to that reported by the UK OpenSAFELY study (Figure 1). Relative to the respective white reference populations, African Americans in the US are at a higher risk compared to the Blacks in the UK. In contrast, Asian people in the US are at a lower risk compared to those in the UK. Further, in the US, the Hispanic population and non-Hispanic American Indian/Alaskan Native population are at substantially elevated risk compared to non-Hispanic white people. External covariate adjustment indicates that accounting for other risk factors, such as various pre-existing conditions that are more prevalent in various minority groups, only explains a small fraction of the racial differences in mortality rates (Extended Data Table 1). These adjusted estimates associated with the age and racial groups, together with estimates of risk associated with the other factors from an underlying fully adjusted model reported in the UK OpenSAFELY study, are used to define risk-score for individuals in the US (Extended Data Table 1).

**Figure 1.**
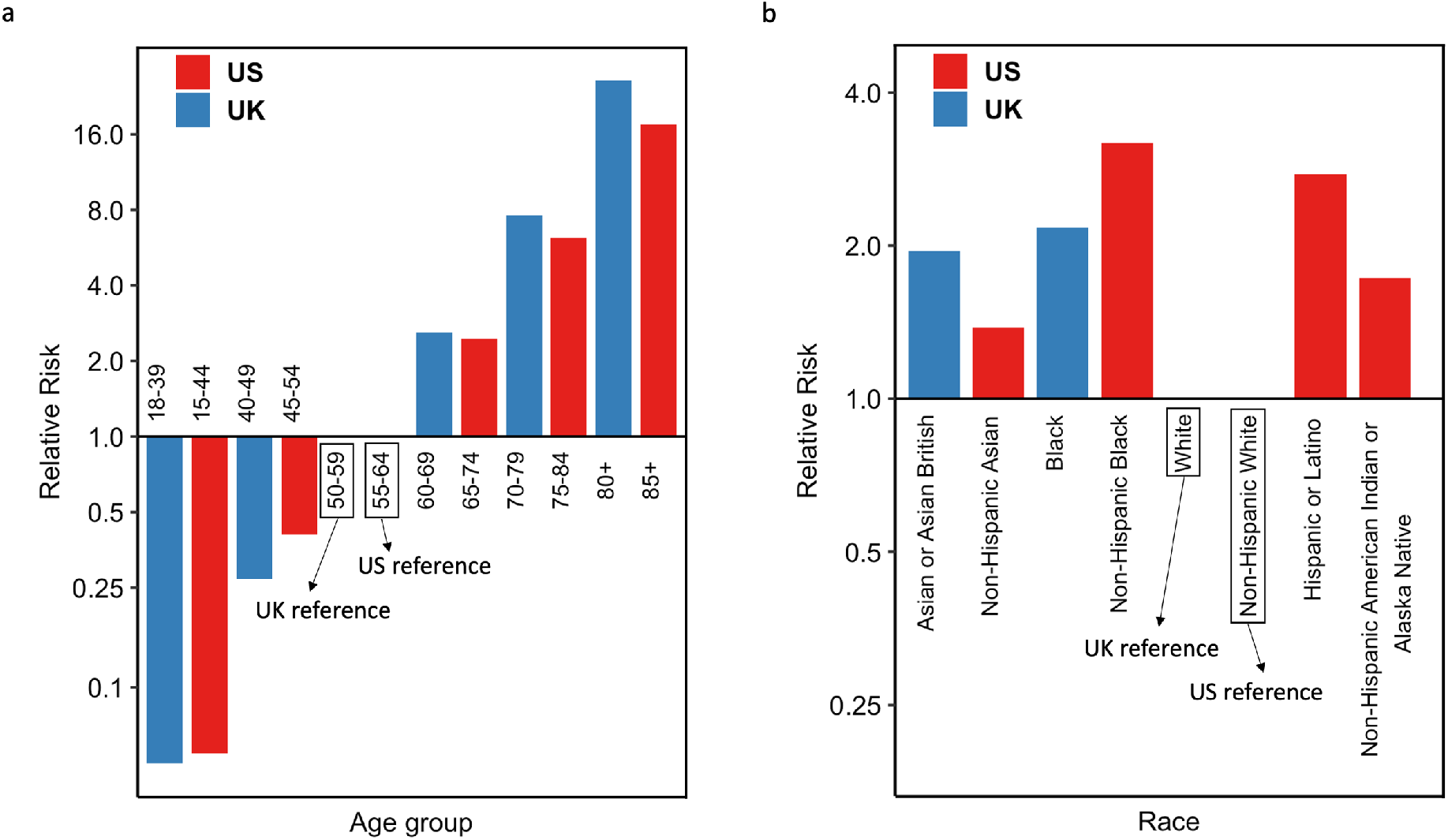
Comparison of COVID-19 mortality risks associated with various age and race/ethnic groups between the US and the UK. **a**, relative risks of various age groups in the UK versus that of in the US. **b**, relative risks of various ethnic groups in the UK versus that of in the US. For UK, the relative risk is from age-sex adjusted model in Table 1 of the UK OpenSAFELY study.^13^ For US, the relative risk associated with race/ethnic groups is adjusted for both age and state whereas the relative risk associated with age is adjusted for state using Poisson regression model fitted to the CDC death count data.^21^

### Risk calculator

We make available a web-based risk-calculator that allows an individual to input information on risk-factors and obtain estimates of individualized risk for COVID-19 mortality in both relative and absolute risk scales (Figure 2). The relative risks for individuals are reported based on the underlying risk-score benchmarked with respect to a “population average risk” defined as population weighted average risk (IER) across the cities. The calculator returns a numerical value for relative-risk and a color-coded categorization of risk into 5 categories. Further, for each person, information on risk-score is combined with projections available from pandemic forecasting models in their state of residence to report an absolute rate of mortality over a specified period of time.

**Figure 2.**
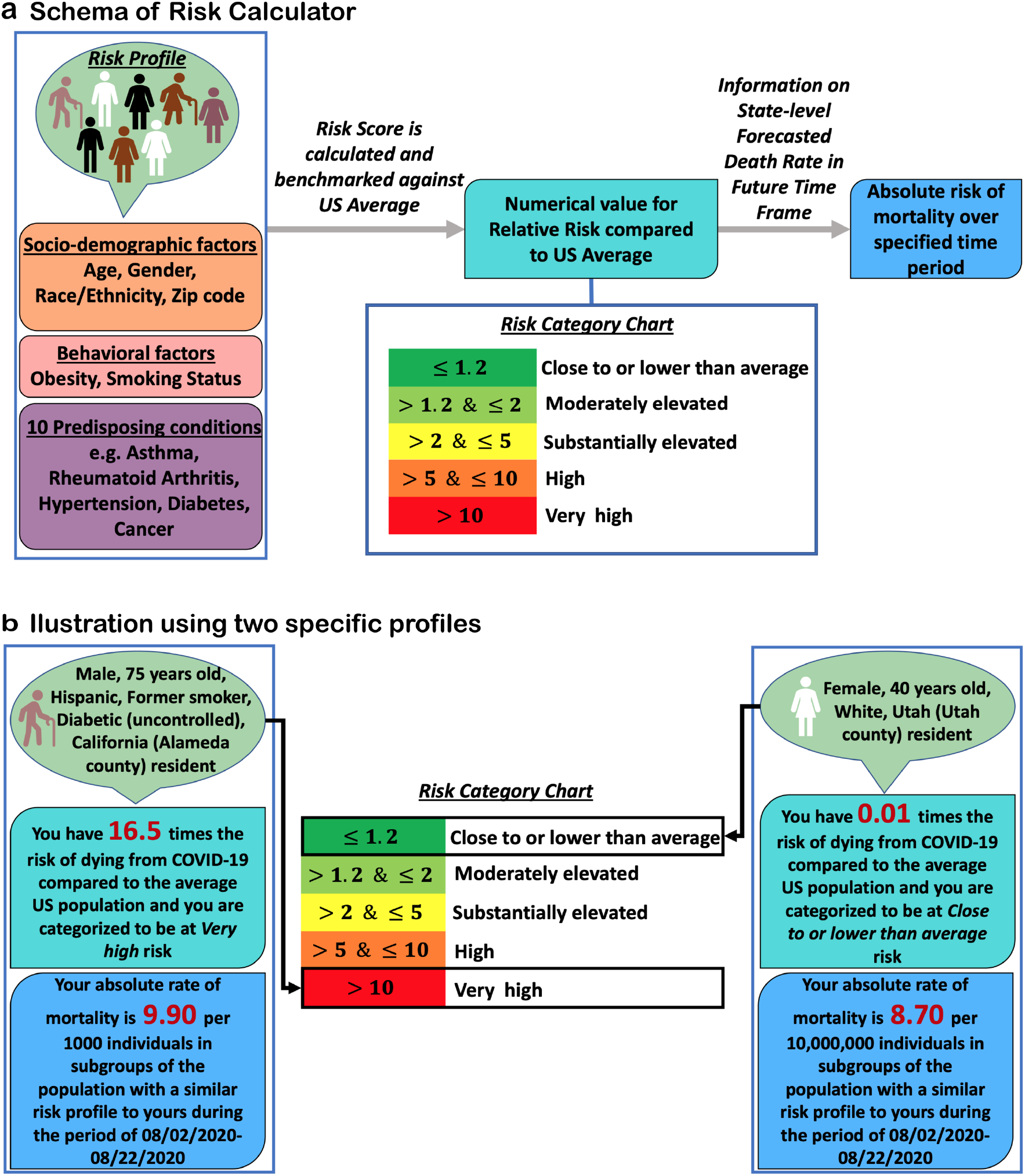
Risk calculator workflow. **a**, the general schema of the risk calculator which takes in the information on socio-demographic, lifestyle and predisposing conditions of an individual to estimate his/her relative risk compared to the average risk in the US adult population (age 18+). Further, based on the projected death rate in the state where the individual resides in, the tool evaluates the individual’s absolute risk of death due to COVID-19 during future time frame. **b**, the output from the risk calculator for two hypothetical profiles.

### Distribution of risk in the US general adult population

We evaluate the risk-score for individuals participating in the NHIS to explore the distribution of risk associated with predisposing factors in the general US adult population. Clearly, there is a very wide variation in risk across individuals in the US (Extended Data Figure 1). Overall, for the US adult population, we estimate that 16.8%, 11.0%, 4.3% and 1.7% of the individuals are at or above risk thresholds associated with elevated (≥ 1.2-fold), substantially elevated (≥2-fold), high (≥5-fold) and very-high (≥10-fold) risk categories, respectively (Table 1). The percentage of the populations exceeding these thresholds varies strongly by age. Only a small fraction (0.1%) of the individuals who are younger than 65 exceed the threshold for high risk. We further examine the distribution of various other risk factors among individuals in the defined high-risk groups for the general population (Extended Data Figures 2-3) and the 65+ year old population (Extended Data Figures 4-5). As expected, male, Hispanic, African Americans, and individuals with obesity and various health conditions are more common in the various high-risk groups compared to the general NHIS population. We observe a similar trend for the 65+ old population.

**Table 1.** The estimated percentages of the NHIS population that exceed various risk thresholds, overall and with the 18-64 or 65+ age group. The age distribution was obtained from the most recent US Census Bureau 2019 data,^29^ and information of the other risk-factors within each age group was obtained from the NHIS.^28^ Risk thresholds are evaluated in reference to the average risk over all subjects.

| Risk category | Overall |  | Age 18 - 64 |  | Age 65+ |  |
| --- | --- | --- | --- | --- | --- | --- |
|  | Population size (%) | % of deaths expected to arise from the risk category | Population size (%) | % of deaths expected to arise from the risk category | Population size (%) | % of deaths expected to arise from the risk category |
| ≥ 10-fold risk | 5.6 M (1.7%) | 30.5% | 30.5 K (0.01%) | 0.2% | 5.5 M (7.9%) | 30.5% |
| ≥ 5-fold risk | 14.0 M (4.3%) | 49.8% | 0.3 M (0.1%) | 1.6% | 13.7 M (19.7%) | 56.5% |
| ≥ 2-fold risk | 36.2 M (11.0%) | 71.2% | 3.3 M (1.3%) | 12.8% | 32.8 M (46.9%) | 85.5% |
| ≥ 1.2-fold risk | 55.3 M (16.8%) | 80.8% | 9.9 M (3.8%) | 28.1% | 45.4 M (65.0%) | 94.0% |

### Projection of the distribution of risk across US cities, counties and states

We observe a wide variation in risk due to predisposing factors across US communities (Figure 3). The IER varies around 10-fold across the cities and counties for the underlying adult- and 65+ year old Medicare-population, respectively (Supplementary Tables S1-S2). A number of major cities, including Detroit, Miami, Baltimore City, New Orleans and Philadelphia, rank very high according to this index. The proportion of individuals crossing various risk thresholds varies even more widely across these communities (Supplementary Tables S1-S2). For example, the percentage of the adult populations in cities which exceed the 5-fold risk threshold varies from 0.5 (Provo, UT) to 11.1 (Detroit, MI). Similarly, the percentage of the 65+ year old Medicare populations which exceed the same threshold varies from <1.5% (multiple counties in CO) to >55.0% (multiple counties in TX). Risk distribution for the 65+ year old Medicare population varies substantially across the states as well (Extended Data Figure 6, Supplementary Table S3). Our projections further show that high-risk groups will be disproportionately enriched for deaths across all the communities (Figure 3, Extended Data Figure 6, Supplementary Tables S1-S2). For example, the ratio of the proportion of deaths that are expected to arise from the ≥5-fold risk group to the proportion of the population targeted at the 5-fold risk threshold ranges in 6.7-35.2 across the US cities (Figure 3, Panel e).

**Figure 3.**
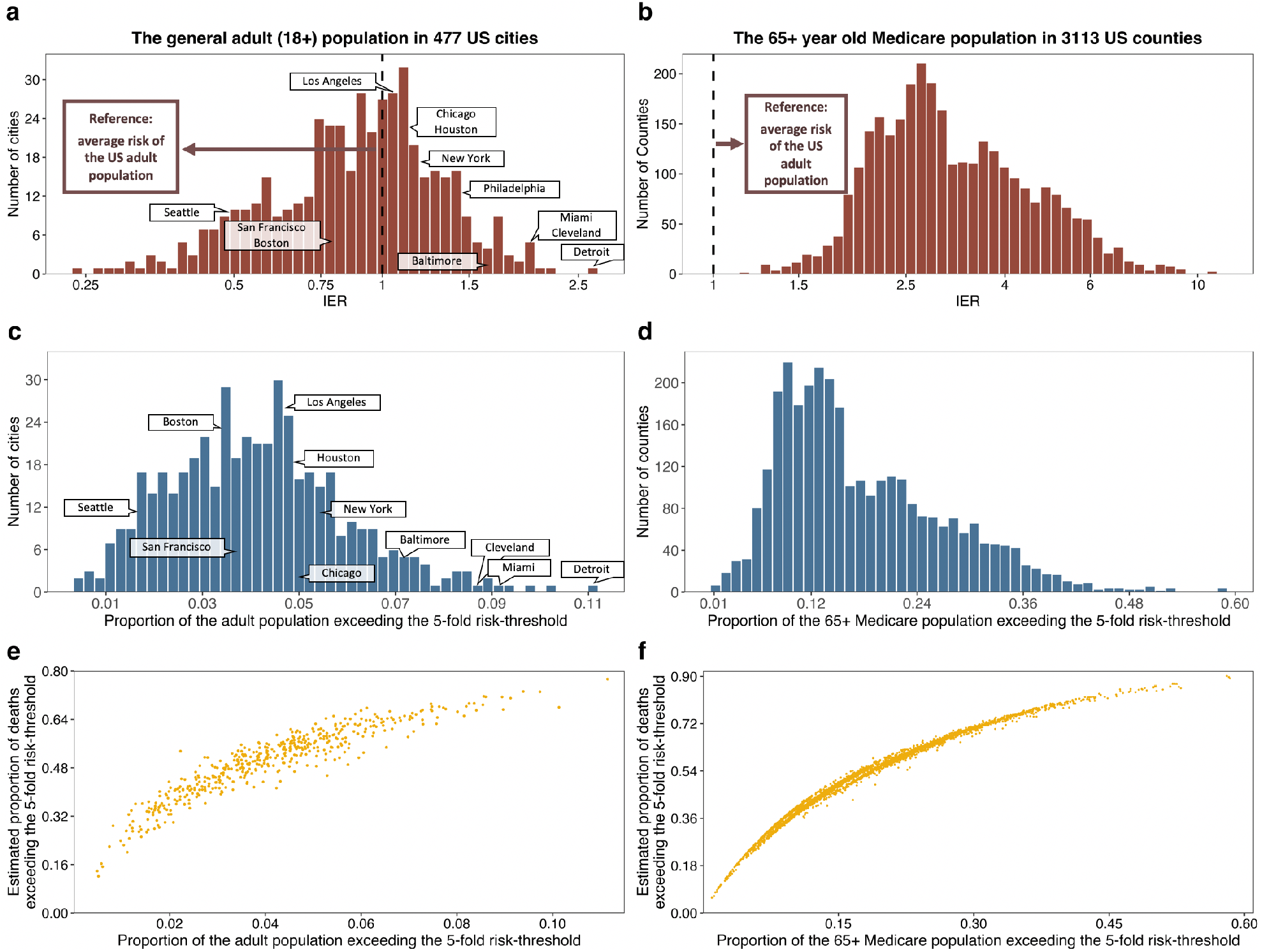
Distribution of the Index of Excess Risk (IER) for COVID-19 mortality and projections for the proportion of high-risk populations (≥5-fold compared to US average risk) across US communities. Left panel: results for the general adult population across 477 US cities; right panel: results for the 65+ Medicare population across 3,113 US counties. **a, b**: the distribution of IER (See Methods and Supplementary Methods, Section 2.3 for the definition of IER). **c, d**: histograms of the proportions of the underlying populations exceeding the 5-fold risk threshold. **e, f**: scatter plots for the proportions of the underlying populations exceeding the 5-fold risk threshold against the proportions of total deaths in the underlying populations that are expected to occur within the ≥ 5-fold risk group. Results for additional risk thresholds are provided in Supplementary Tables S1-S2.

### An independent validation of risk projections using recent deaths in the US

In a negative binomial regression analysis of recent deaths (between June 7 and August 1) in the cities over a moving window of two-weeks, we find the coefficient of log(IER) to be statistically highly significant throughout, with an average value of 0.96 (Supplementary Table S4), close to its ideal value of 1.0, indicating excellent calibration of the underlying individual-level model for the population (see Methods). Further, in a weighted least squares analysis, we find that log(IER) explains on average 15.9% of the variation of death rates (in the logarithmic scale) over this time period across the underlying counties (Figure 4). In comparison, population density and reported infection rate three-week prior explain on average 1.7% and 12.5% of the variance of the underlying death rates, respectively. In a conditional analysis that accounts for major regional differences in pandemic dynamics, we find that IER explains as much or more of the variance of death rates as three-week prior infection rates.

**Figure 4.**
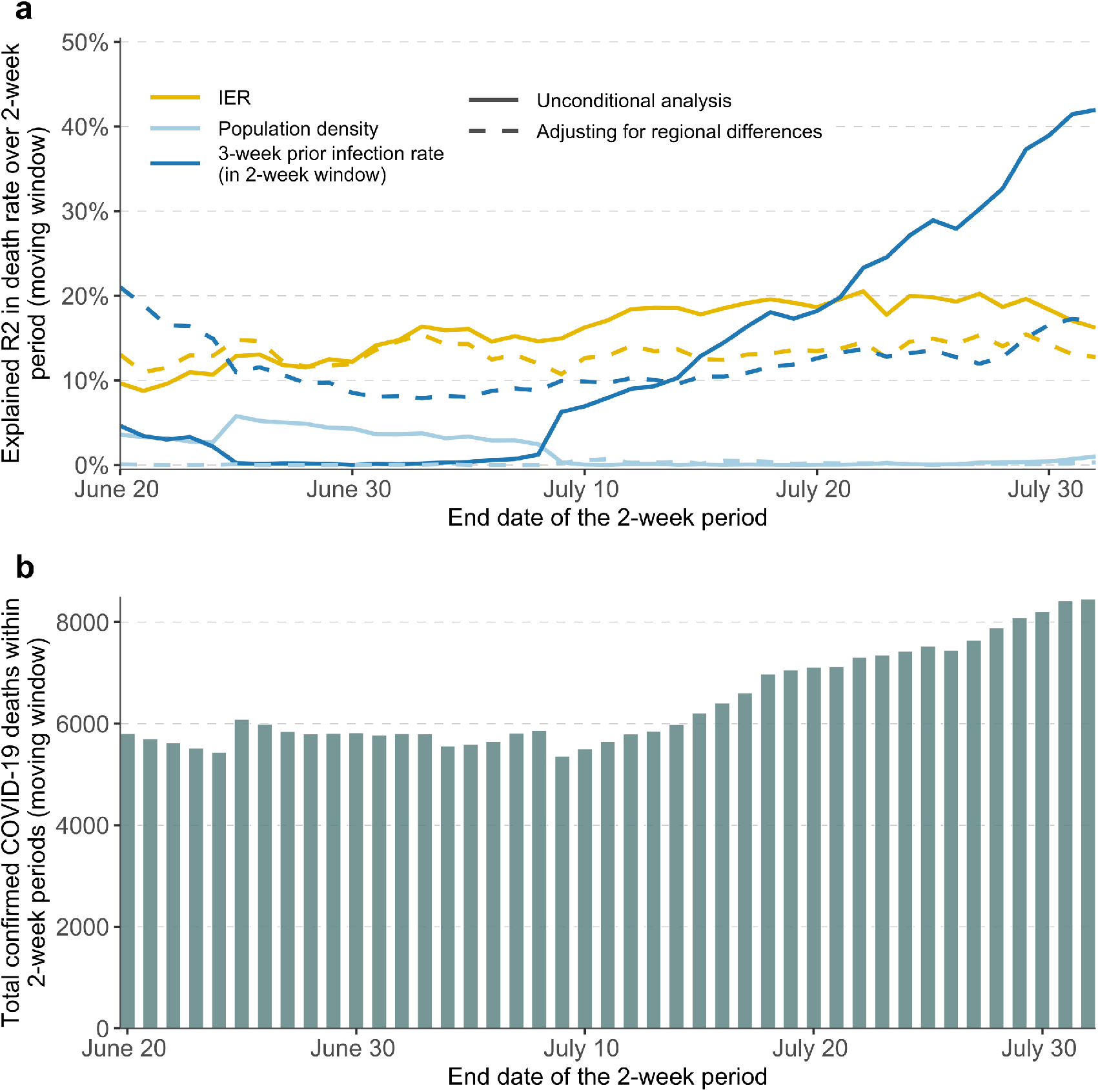
Validation of the risk model using recent deaths (June 7 - Aug 1, 2020) in 477 US cities. **a**, the explained variance (R^2^) of logarithm of death rate in a moving window of two-week periods by IER, population density and three-week prior infection rate (also aggregated over two-week window), all transformed in logarithmic scale. **b**, the cumulative number of confirmed COVID-19 deaths during the corresponding two-week periods. 256 counties that contain the 477 studied cities were included in the validation, where the county-level IER was calculated as the weighted IER with each city being weighted by its population size. During each two-week period only the counties with non-zero deaths and infections were included in the analyses.

## Discussion

In this article, we have developed a risk calculator for COVID-19 mortality for the general US population by combining multiple data sources. The tool is unique in the regard that it combines information from individual-level risk factors, as well as community-level risks, to project the absolute rate of mortality for different risk profiles. We have further applied this tool to data available from various national databases to identify high-risk cities/counties and estimate the size of the populations at various levels of risks within these communities. Our tools and results could inform policy developments for equitable distribution of early vaccines and other scarce preventive resources.

Promising preliminary results on immunogenicity and safety from Phase-I/II of a number of recent trials^18,19^ have raised the hope that vaccines could be ready for distribution by the end of this year. However, production of vaccines for wide distribution at a national and global scale will take months and years. In recognizing that vaccines, when becoming available, will be constrained by supply, a number of national and international bodies^20,35^ have called for the development of frameworks that will allow equitable distribution of vaccines, taking into account differential risks for individuals and communities associated with various factors including age, racial disparity, socioeconomic conditions, high-risk occupations, pre-existing conditions and community level risk for infection. Specifically, it has been noted that the allocation of scarce therapeutics and vaccines should be guided by “size, distribution and risk-profiles of the affected population”.^36^

In this article, we have provided a detailed framework for developing risk tools by combining multiple data and information sources, and in using such tools to make projections of risks at both individual and population levels. We have shown how a risk tool can be applied to various population-based databases to estimate the size of high-risk populations at the community level that could be gradually prioritized for vaccination. Further, our framework allows evaluation of absolute risks taking into account population-level pandemic dynamics in a given time frame, and thus could be used to refine guidelines if the pandemic surges or ebbs disproportionately in certain regions. While we plan to extend the model in the future by incorporating additional risk-factors such as occupational exposures, we believe the estimates we currently provide for the size of high-risk populations for the cities, counties, states and US overall could be immediately useful for national and local policymakers in planning for vaccine distribution around the country. Currently, the Advisory Committee on Immunization Practices (ACIP) of the US CDC has developed a broad 5-tier plan for vaccine distribution starting first with health care workers,^37-39^ but it will be imperative that these guidelines are refined based on the actual level of risk of individuals within and across different priority groups.

In principle, a similar framework can also be useful for developing strategies for the global distribution of vaccines and preventive therapeutics. One could define various thresholds based on absolute risks that would be applied uniformly across countries to decide which individuals should be prioritized at various phases of vaccine dissemination. Then the projection tools we have developed can be applied to any available country-specific data on distribution of underlying riskfactors to estimate the size of the target populations and accordingly available vaccines can be allocated across countries. Currently, we are working with investigators from the Pan American Health Organization (PAHO) to identify such datasets and provide estimates of the size of various high-risk populations across countries in South America. The development of policy for efficient and impactful distribution of vaccines, however, will also depend on many other factors, including, but not limited to cost, overall social benefit, implementation issues and available infrastructures.

Our risk tools and projections can also be useful for identifying high-risk groups who would benefit most from “shielding” efforts until they can be vaccinated. In the beginning of the pandemic, the National Health Service of the UK identified about 1.5 million individuals to be at extremely high risk due to selected conditions and provided them with assistance for food delivery and medical services.^40^ In California, local and state government developed the Project Roomkey^41^ to provide free hotel rooms, meals and other services to asymptomatic homeless people who are at high risk due to their age or/and underlying conditions. In the future, as businesses, schools, and higher education institutes consider reopening, strategies need to be in place to identify and shield highrisk individuals. Finally, general population risk tools can also help individuals understand future risk for serious outcomes, not only for themselves, but also for family members and friends, and thus could better motivate them to adhere to standard guidelines for infection prevention, such as handwashing and mask wearing.

A few studies in the past have investigated the proportions of “high-risk” individuals for COVID-19 related serious illness or mortality in the UK, the US and across nations globally.^40,42-44^ These studies have defined high-risk individuals based on prevalence of one or more risk factors without taking into account the relative contribution of these factors. Further, because of the broad definition used, they estimate that a very large fraction of the populations, 20% in UK and 16-31% globally, are at “high risk”. In contrast, we have defined different risk categories based on an underlying score that allows one to assign a more precise magnitude of risks to these categories. Further, our framework allows evaluation of future absolute risks for individuals and communities, incorporating information from pandemic forecasting models, and thus is uniquely suitable for planning vaccination and other prevention efforts across regions that may have wide variation in the infection dynamics.

Our studies have several limitations. First, information on risk for the majority of the risk factors was derived from the UK-based OpenSAFELY study. However, we modified the model to make it more suitable for the US population by incorporating population-based information on age and race associated rate of mortality and by performing external covariate adjustment to account for their correlation with other risk-factors. Further, we have empirically shown through independent validation analyses that the projected risks are well calibrated for the general US population and correlate strongly with recent death rates across counties in the US. There is, however, an urgent need for individual-level data from large population-based studies, akin to the UK OpenSAFELY study, with detailed information on both outcomes and risk factors in the US setting. US-specific data are particularly needed to understand the risk associated with measures of social deprivation which have been shown to be an important risk factor independent of race/ethnicity and pre-existing conditions.

Another limitation of our current tools and projections is that they do not incorporate information associated with front line occupations that clearly pose higher risks for infection. The Office of National Statistics (ONS)^45^ in the UK has released data identifying several frontline occupations that are at increased risk of COVID-19 mortality. We have mapped these occupation categories in the NHIS dataset and have observed that various minority populations are over-represented in these groups (Supplementary Table S5). We, however, have not incorporated this risk information available from the UK to our risk tool as the level of virus exposures the individuals with these occupations received in the US may be different.^46^ In the future, we will explore data for validation of or/and re-estimation of risk of mortality associated with various occupation categories in the US setting.

In summary, we present a comprehensive and rigorous framework, and a set of associated tools, for assessing general population risks of COVID-19 mortality incorporating individual profiles, distributions of various risk-factors and population-level pandemic dynamics. Our risk projection results for the US cities/counties could be directly used for guiding strategies for equitable allocation of early vaccines and other preventive resources in the coming months. Further, our risk tool and the underlying statistical methodologies can be applied to carry out similar analyses internationally and thus inform prevention efforts globally.

## Data Availability

All data used in the manuscript are publically available and can be accessed at https://github.com/nchatterjeelab/COVID19Risk/tree/master/data. All codes for data management and analysis can be accessed at https://github.com/nchatterjeelab/COVID19Risk.

## Web resources

The web-based tool for individualized risk calculator and the interactive maps for viewing the city, county, state and national level risk projections in the US are at http://covid19risktools.com/.

## Data availability

All data used in the manuscript are publicly available and can be accessed at https://github.com/nchatterjeelab/COVID19Risk/tree/master/data.

## Code availability

The R codes for data management and analyses in this article can be accessed at https://github.com/nchatterjeelab/COVID19Risk.

## Acknowledgements

We thank Dr. Allison Meisner from the Johns Hopkins University, Biostatistics Department and Dr. Montserrat García-Closas from Division of Cancer Epidemiology and Genetics at National Cancer Center for their comments on a previous version of the manuscript. The funding for this research came from the Bloomberg Distinguished Professorship endowment.

## Author contributions

J.J., N.A., P.K. and N.C developed all methods. J.J, N.A, P.K and Y.Z. conducted the data analyses. B.H. developed the web-based tool with assistance from J.J., N.A. and P.K. E.W. developed the interactive maps. N.C., J.J., N.A. and P.K. wrote the first draft of the manuscript. All authors reviewed the final manuscript.

---

**Extended Data Figure 1.**
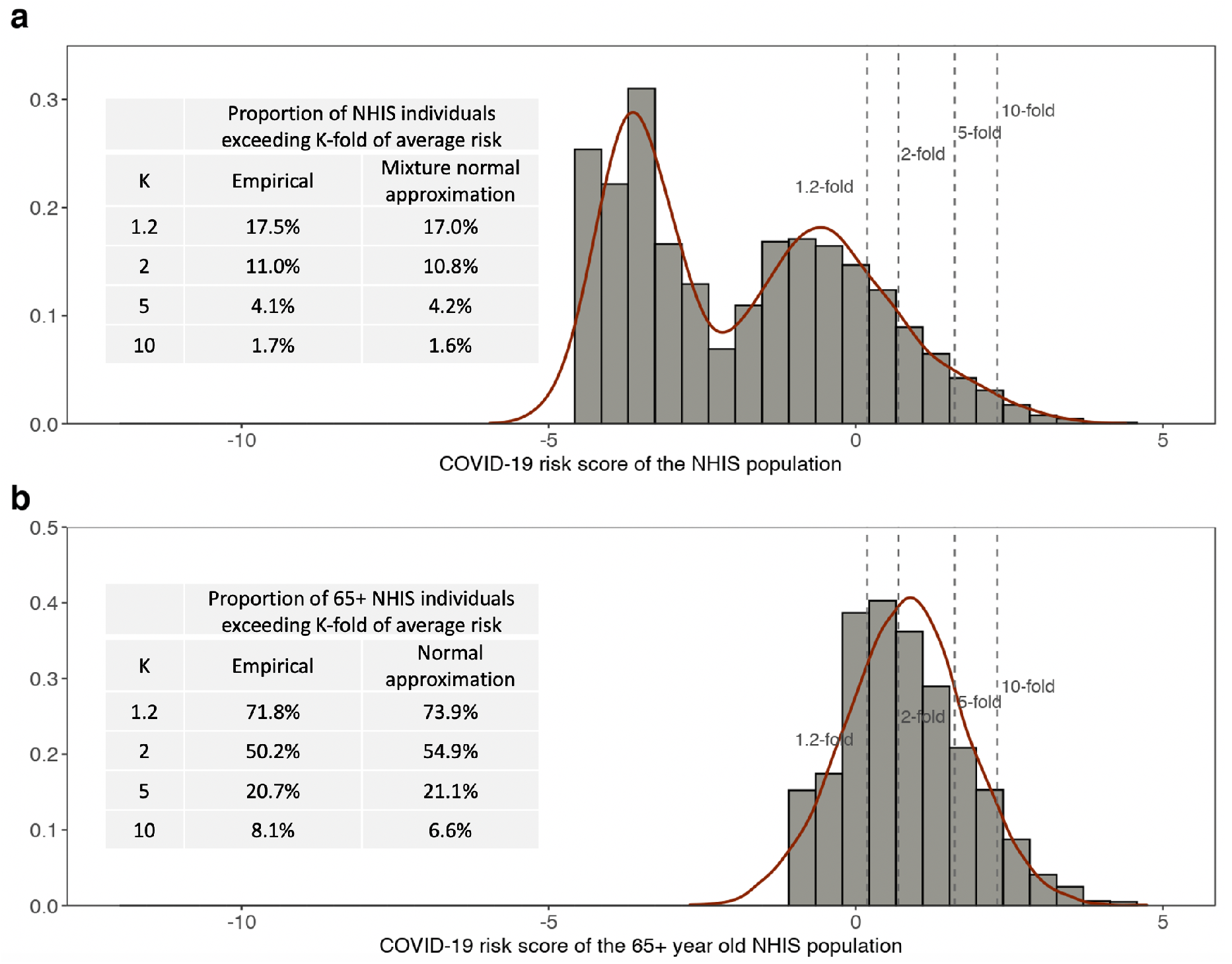
Distribution of risk-score across the general NHIS population (18+ year old, a) and across the 65+ year old NHIS population (b). Empirical distributions are compared with those based on mixture-normal or normal approximations. The risk-score is calculated based on age, gender, ethnicity and 12 different health conditions, but not social deprivation index (SDI) due to the absence of the relevant data in NHIS. The risk-score is centered using a reference value that corresponds to the average risk across the individuals in NHIS.

**Extended Data Figure 2.**
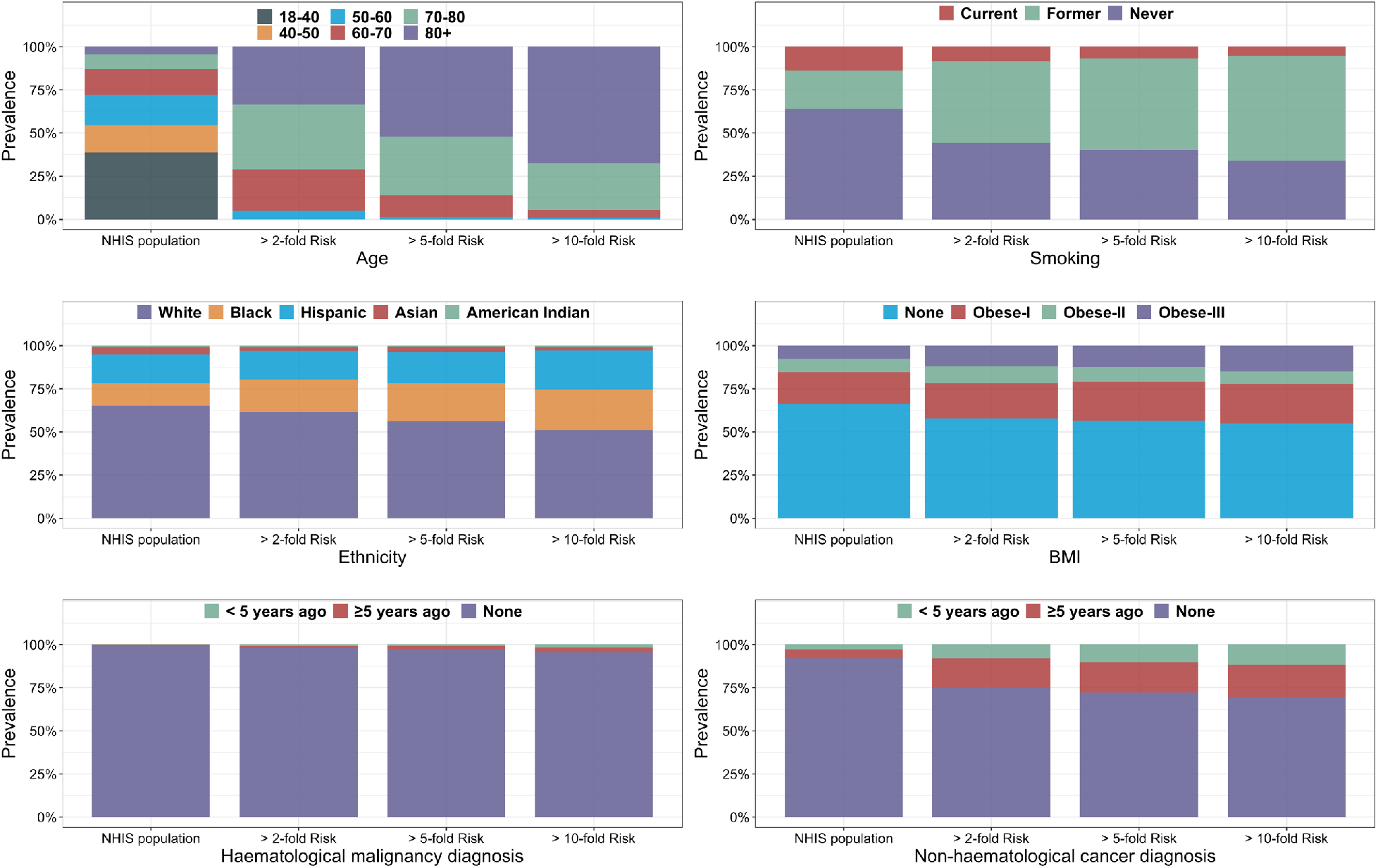
Distribution of risk factors (age, smoking status, ethnicity, BMI, hematological and non-hematological cancer diagnosis) in the general NHIS population and among individuals in different risk groups. The risk score is calculated based on the demographic covariates and 12 different health conditions, excluding SDI which is unavailable in NHIS. The risk thresholds are defined with respect to the average risk of the NHIS population.

**Extended Data Figure 3.**
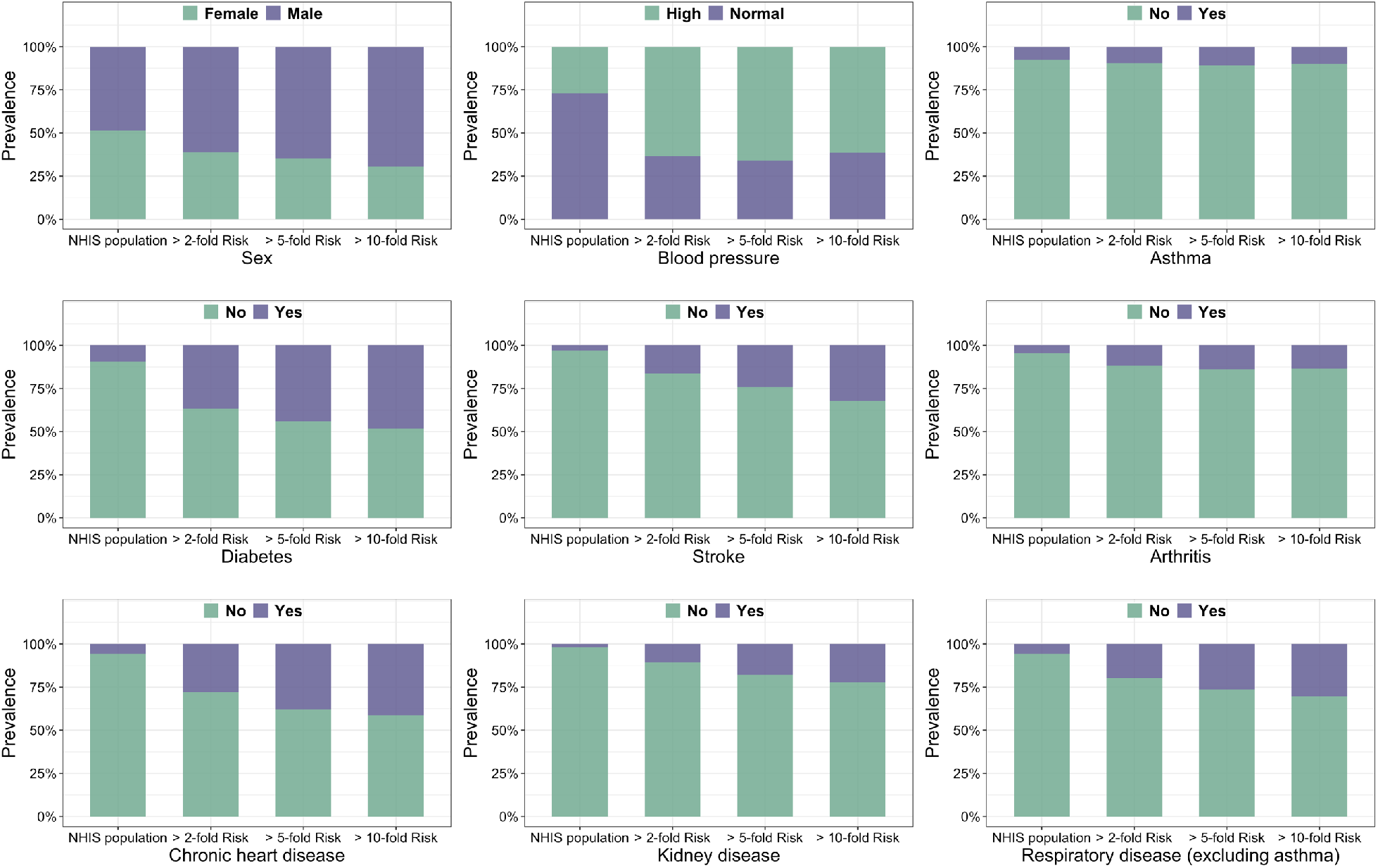
Distribution of risk factors (sex, blood pressure, status of asthma, diabetes, stroke, arthritis, chronic heart disease, kidney disease and respiratory disease excluding asthma) in the general NHIS population and among individuals in different risk groups. The risk score is calculated based on the demographic covariates and 12 different health conditions, excluding SDI which is unavailable in NHIS. The risk thresholds are defined with respect to the average risk of the NHIS population.

**Extended Data Figure 4.**
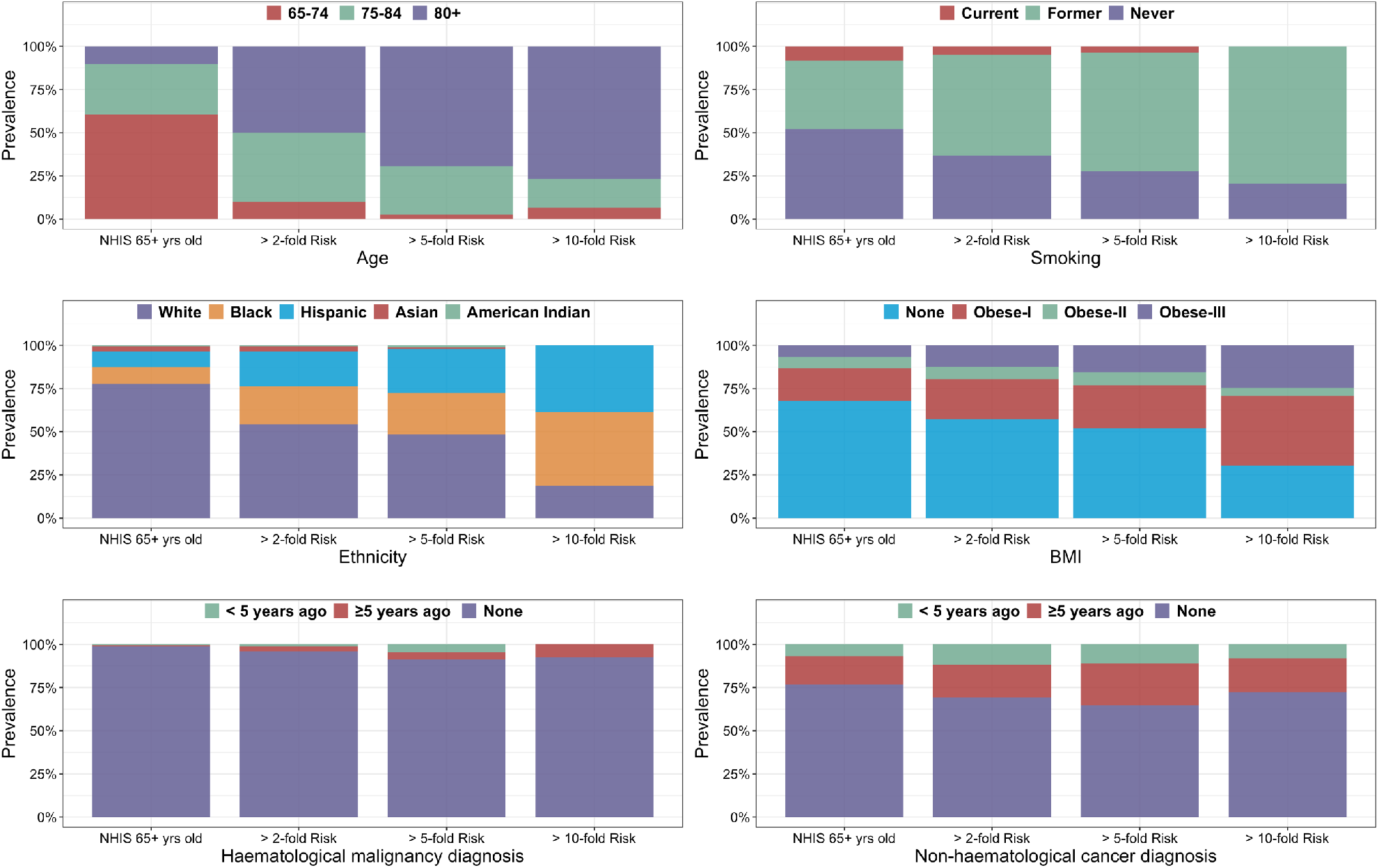
Distribution of risk factors (age, smoking status, ethnicity, BMI, hematological and non-hematological cancer diagnosis) in the 65+ years old NHIS and among individuals in different risk groups. The risk score is calculated based on the demographic covariates and 12 different health conditions, excluding SDI which is unavailable in NHIS. The risk thresholds are defined with respect to the average risk of the NHIS population.

**Extended Data Figure 5.**
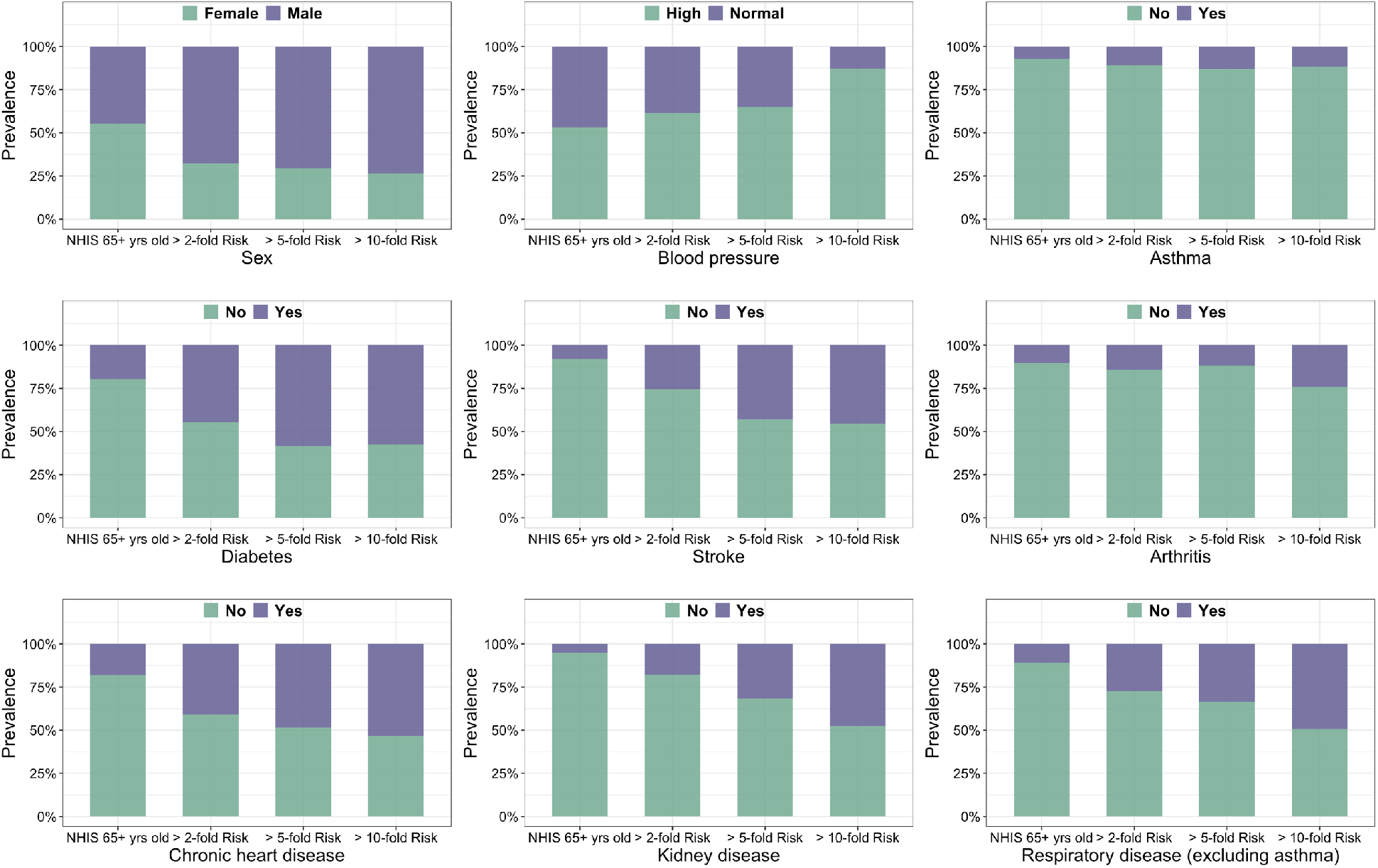
Distribution of risk factors (sex, blood pressure, status of asthma, diabetes, stroke, arthritis, chronic heart disease, kidney disease and respiratory disease excluding asthma) in the 65+ years old NHIS population and among individuals in different risk groups. The risk score is calculated based on the demographic covariates and 12 different health conditions, excluding SDI which is unavailable in NHIS. The risk thresholds are defined with respect to the average risk of the NHIS population.

**Extended Data Figure 6.**
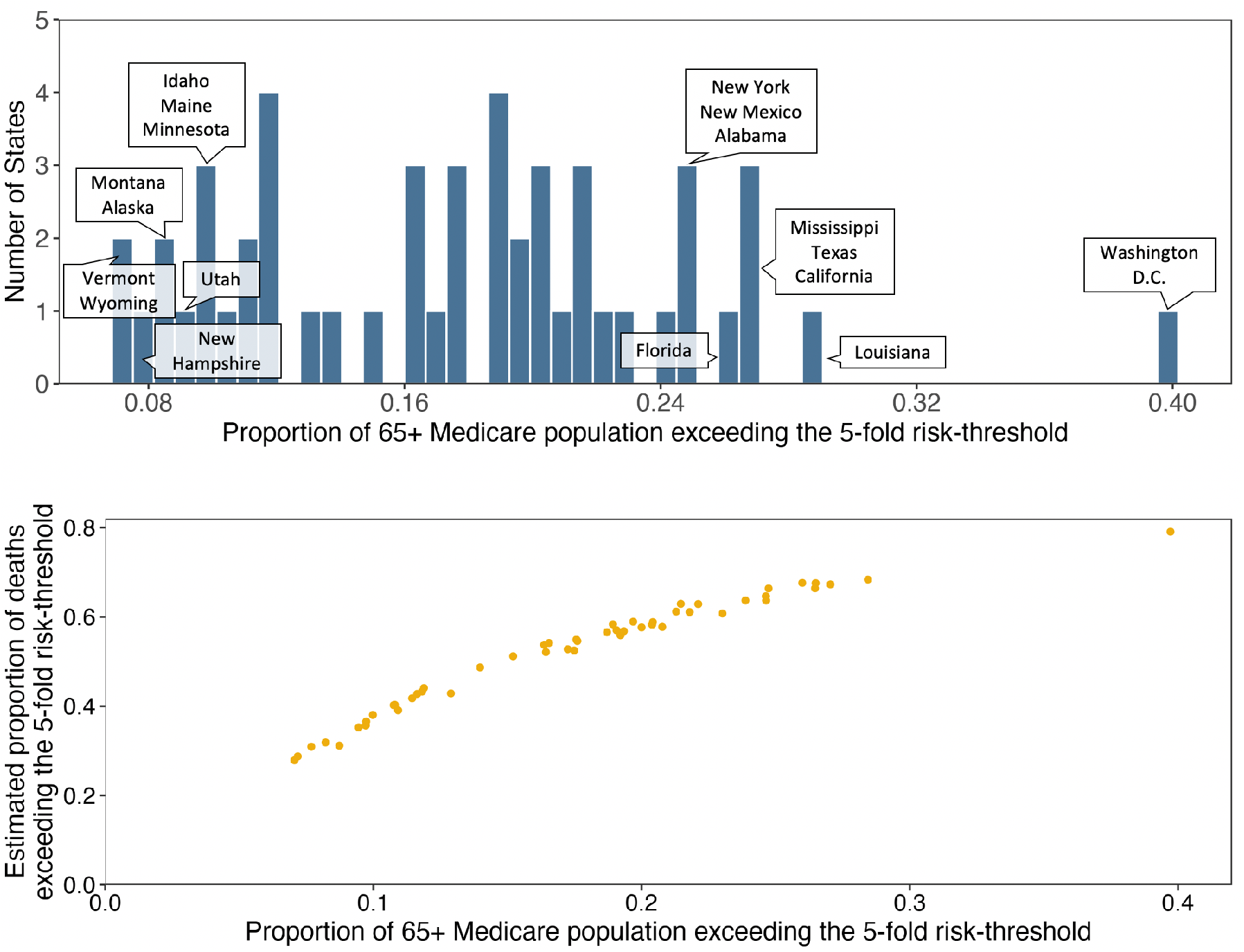
Projections for high-risk (≥5-fold risk compared to the US average) for the 65+ year old Medicare population across US states. **a**, histogram of the proportion of population exceeding the 5-fold risk threshold across states. **b**, scatter plot of the proportion of population exceeding the 5-fold risk threshold against the proportion of deaths among the population that are expected to occur within the ≥5-fold risk group. Results for additional risk thresholds are provided in Supplementary Table S3.

**Extended Data Table 1.** Model coefficient (log hazard ratio of COVID-19 death) for each
category of the various risk factors.

| Variable type | Variables | Log-hazard ratio |  |  | Notes |
| --- | --- | --- | --- | --- | --- |
|  |  | UK OpenSAFELY | US CDC | Fully adjusted |  |
| Socio-demographic | **Age** |  |  |  |  |
|  | 15-<45 | - | -2.996 | -3.037 | Assumed the estimate for 18-<45 age group is the same as this group |
|  | 45-<55 | - | -0.968 | -0.994 |  |
|  | 55-<65 | - | 0 | 0 | Reference category |
|  | 65-<75 | - | 0.952 | 0.751 |  |
|  | 75-<85 | - | 1.898 | 1.613 |  |
|  | 85+ | - | 2.997 | 2.733 |  |
|  | **Race/Ethnicity** |  |  |  |  |
|  | White | - | 0 | 0 | Reference category |
|  | Black | - | 1.157 | 0.948 |  |
|  | Hispanic | - | 1.015 | 0.833 |  |
|  | Asian | - | 0.322 | 0.300 |  |
|  | American Indian/Alaska native | - | 0.542 | 0.536 |  |
|  | **IMD quintile** |  |  |  |  |
|  | 1 (least deprived) | 0 | - | 0 | Reference category |
|  | 2 | 0.174 | - | 0.174 |  |
|  | 3 | 0.231 | - | 0.231 |  |
|  | 4 | 0.425 | - | 0.425 |  |
|  | 5 (most deprived) | 0.531 | - | 0.531 |  |
|  | **Sex** |  |  |  |  |
|  | Female | 0 | - | 0 | Reference category |
|  | Male | 0.657 | - | 0.657 |  |
| Behavioral | **BMI** |  |  |  |  |
|  | Not Obese | 0 | - | 0 |  |
|  | Obese class I | 0.247 | - | 0.247 |  |
|  | Obese class II | 0.470 | - | 0.470 |  |
|  | Obese class III | 0.824 | - | 0.824 |  |
|  | **Smoking status** |  |  |  |  |
|  | Never | 0 | - | 0 | Reference category |
|  | Ex-smoker | 0.278 | - | 0.278 |  |
|  | Current | -0.062 | - | -0.062 |  |
| Predisposing conditions | **Blood pressure** |  |  |  |  |
|  | Normal | 0 | - | 0 | Reference category |
|  | Hypertension | -0.030 | - | -0.030 |  |
|  | Respiratory disease excluding Asthma | 0.582 | - | 0.582 |  |
|  | Asthma | 0.049 | - | 0.049 |  |
|  | Chronic heart disease | 0.239 | - | 0.239 |  |
|  | **Diabetes** |  |  |  |  |
|  | Controlled | 0.385 | - | 0.385 |  |
|  | Uncontrolled | 0.802 | - | 0.802 |  |
|  | **Non-hematological cancer** |  |  |  |  |
|  | <1 year | 0.519 | - | 0.519 |  |
|  | 1-5 year | 0.191 | - | 0.191 |  |
|  | ≥5 year | 0.020 | - | 0.020 |  |
|  | **Hematological cancer** |  |  |  |  |
|  | <1 year | 1.194 | - | 1.194 |  |
|  | 1 - 5 year | 1.230 | - | 1.230 |  |
|  | ≥5 year | 0.610 | - | 0.610 |  |
|  | Stroke/Dementia | 0.560 | - | 0.560 |  |
|  | Kidney disease | 0.565 | - | 0.565 |  |
|  | Rheumatoid arthritis | 0.157 | - | 0.157 |  |

